# Performing risk stratification for COVID-19 when individual level data is not available – the experience of a large healthcare organization

**DOI:** 10.1101/2020.04.23.20076976

**Authors:** Noam Barda, Dan Riesel, Amichay Akriv, Joseph Levi, Uriah Finkel, Gal Yona, Daniel Greenfeld, Shimon Sheiba, Jonathan Somer, Eitan Bachmat, Guy N. Rothblum, Uri Shalit, Doron Netzer, Ran Balicer, Noa Dagan

## Abstract

With the global coronavirus disease 2019 (COVID-19) pandemic, there is an urgent need for risk stratification tools to support prevention and treatment decisions. The Centers for Disease Control and Prevention (CDC) listed several criteria that define high-risk individuals, but multivariable prediction models may allow for a more accurate and granular risk evaluation. In the early days of the pandemic, when individual level data required for training prediction models was not available, a large healthcare organization developed a prediction model for supporting its COVID-19 policy using a hybrid strategy. The model was constructed on a baseline predictor to rank patients according to their risk for severe respiratory infection or sepsis (trained using over one-million patient records) and was then post-processed to calibrate the predictions to reported COVID-19 case fatality rates. Since its deployment in mid-March, this predictor was integrated into many decision-processes in the organization that involved allocating limited resources. With the accumulation of enough COVID-19 patients, the predictor was validated for its accuracy in predicting COVID-19 mortality among all COVID-19 cases in the organization (3,176, 3.1% death rate). The predictor was found to have good discrimination, with an area under the receiver-operating characteristics curve of 0.942. Calibration was also good, with a marked improvement compared to the calibration of the baseline model when evaluated for the COVID-19 mortality outcome. While the CDC criteria identify 41% of the population as high-risk with a resulting sensitivity of 97%, a 5% absolute risk cutoff by the model tags only 14% to be at high-risk while still achieving a sensitivity of 90%. To summarize, we found that even in the midst of a pandemic, shrouded in epidemiologic “fog of war” and with no individual level data, it was possible to provide a useful predictor with good discrimination and calibration.

## Introduction

The global coronavirus disease 2019 (COVID-19) pandemic is challenging healthcare systems around the world^1,2^. The wide range of outcomes observed, ranging from subpopulations that are mainly asymptomatic to subpopulations with substantial fatality rates^3^, calls for risk stratification.

Such risk stratification can help to better tailor efforts, both for prevention (i.e. home quarantine, social distancing) and for treatment of confirmed cases (i.e. hospitalization vs. community isolation). In the face of equipment, services and personnel shortages, risk stratification can allow better use of existing resources and improved outcomes.

To define who is at high risk for severe disease, the Centers for Disease Control and Prevention (CDC) defined the following criteria^4^: people 65 years and older, people who live in nursing homes, and people with at least one of the following conditions – chronic lung disease, serious heart conditions, severe obesity, diabetes, chronic kidney disease, liver disease or people who are immunocompromised.

Unlike the binary classification to low and high risk groups that results from decision rules such as the ones put forth by the CDC, the medical community is long accustomed to use more granular and individualized evaluation of patients’ risk, which are usually quantified using multivariable prediction models^5^. Because patient risk is multi-factorial in nature, with many interactions between the various factors, such models are well suited to the task of risk evaluation. Training of these models requires individual level data, which is usually available from retrospective electronic healthcare record (EHR) data^6,7^ or from datasets of cohorts that were collected for research purposes^8,9^. Such individual level data of COVID-19 patients is starting to accumulate in different countries, but was not available when most western countries started forming their strategy to deal with the rise in COVID-19 patients.

Many attempts to provide risk prediction models for COVID-19 were described in recent weeks. Wynants et. al recently reviewed^10^ prediction models for COVID-19, and found three that were trained on a general population to predict hospital admission due to pneumonia (as a proxy for COVID-19 pneumonia), and ten other prognostic models that were trained on COVID-19 patients to predict an outcome of death, severe disease or the need for hospital admission. However, most of these models are not reported at the usual standard required for prediction models^11^, and Wynants et. al summarized them as “poorly reported, at high risk of bias, and their reported performance is probably optimistic”.

In this paper we propose a hybrid methodology, which allows the construction of a multivariable prediction model without access to individual level data pertaining to the current pandemic. This methodology first uses a baseline model trained on the general population in order to provide a granular ranking (discrimination) of the risk for severe respiratory infection or sepsis, which were hypothesized to share a common physiologic tendency with severe COVID-19 infection. Then, a post-processing multi-calibration algorithm^12^ is used to adjust the predictions to published aggregate epidemiological reports of COVID-19 case fatality rates (CFRs) in various subpopulations.

This methodology was applied in practice during the first weeks of the COVID-19 pandemic. The resulting model was then used for different purposes within a large integrated healthcare organization. Once sufficient individual level data for COVID-19 became available, the model was validated.

## Methods

### Setting and source of data

This is a retrospective cohort study based on the data warehouse of Clalit Health Services (CHS), a large integrated payer-provider operating in Israel. Health insurance in Israel is mandatory and is offered by several payer-provider organizations that directly supply most of the medical services to their insured population and indirectly pay for additional required services. CHS, the largest of these organizations, insures over half of the Israeli population (approximately 4.5 million members), which are a representative sample of the entire population. The CHS data warehouse contains both medical data (primary care, specialist care, laboratory data, in-network hospitalization data, imaging data etc.) and claims data (mostly out-of-network hospitalization data). CHS has been digitalized since the year 2000 and has a low yearly attrition rate (∼1-2%), allowing long follow-up of patients in the data warehouse.

### Baseline Model

The entire CHS population over the age of ten years was designated as relevant for the prediction. The exclusion of patients under the age of ten years was done based on the very low rates of COVID-19 mortality described in this young population, in contrast to young children’s susceptibility to severe respiratory infections by other pathogens (which may have been detected by the baseline model and would potentially create bias in the final COVID-19 model).

The index date was set to June 1^st^, 2018. Baseline covariates were extracted in the year prior to this date, with the value set as the last result available. A positive outcome was defined as a hospitalization with diagnoses of pneumonia, other respiratory infections or sepsis, or a positive influenza PCR result (i.e. influenza infection that required inpatient care). Outcome data was collected over a follow-up period of 12 months (until May 31^st^, 2019).

The baseline model was trained in two phases – initial training for feature selection and final training for creation of the baseline model. In the first phase, a population of 1,000,000 members, selected randomly from CHS members of the relevant age ranges, was selected. This population was run through an automatic prediction pipeline to explore important features for the prediction task, out of all covariates available in the CHS data warehouse (≈15,000 potential features).

Once this model was trained, the top 30 features were passed to a group of clinicians that chose a subset of features with meaningful and intuitive medical sense. In addition, variables describing medical conditions reported to be related to severe COVID-19 infection were added manually. This selection process, which resulted in 24 selected features, was performed to improve the interpretability of the resulting model and to allow its sharing with other providers. The full list of features used in the eventual model and the exact definition of the outcome used are detailed in Supplementary Table 1.

The final baseline model, with the selected features, was trained and tested using another random sample of 1,050,000 individuals (that were not included in the feature selection phase). This population was then randomly divided in a 70/30 ratio to training-validation and test sets, respectively.

The final model employed was a decision-tree-based gradient boosting model using the LightGBM library^13^ with default hyperparameters. The validation set was used for early stopping, with area under the receiver operating characteristics curve (AUROC) used as the performance measure. Decision tree models are capable of handling missing data without the need for imputation, and accordingly, none was performed.

The model was scored on the test set for metrics of discrimination and calibration. Discrimination was measured as the AUROC. Calibration was measured using a smooth calibration plot^14^. Contribution and effect of the selected features was presented using SHAP scores^15^. Confidence intervals (CI) for the various performance measures were derived using the bootstrap percentile method^16^ with 5,000 bootstrap repetitions.

### Adjustment of the baseline model predictions for COVID-19 mortality

The final baseline model was applied to the entire CHS population over the age of 10 years as of an index date of February 1^st^, 2020, which was chosen to avoid the effects of COVID-19 disease (the first COVID-19 case was identified in Israel at January 29^th^, 2020). After producing individual risk assessment for what we hypothesized to be a physiologic tendency to severe COVID-19 infection, there was a need to recalibrate the results to the outcome of interest – mortality due to COVID-19 infection. This process was done using reported COVID-19 CFRs for different subpopulations, taken from a report by the Chinese center for disease control^17^. The data in this report was presented as one-way conditionals, e.g., the probability of mortality given being in a specific age range, or of a specific sex group.

Before applying the one-way conditionals from the Chinese population to the Israeli population, they were corrected to account for the different demographic distributions of the two populations. For this purpose, a linear probability model was trained on the Chinese data with ten-year age groups and sex as the predictors. This model has the advantage of only requiring the pair-wise covariance between the independent variables, which were available for the Chinese population as part of the global burden of disease (GBD) study 2017^18,19^. Once this model was trained, the corresponding conditionals were calculated on the Israeli population, using its demographic characteristics.

Adjustment of the predictions from the baseline model was done using a multi-calibration algorithm^12^. This algorithm acts as a post-processing phase for any baseline prediction model and works by iteratively adjusting the predictions of each subgroup (e.g., females aged 60-69) so that their mean is equal to the mean of the observed outcomes in that subgroup. The algorithm iterates until no subgroups remain whose average is further from their target value by more than a pre-determined tolerance, which was set to 1%. Since the subpopulations are overlapping, this process requires several iterations until it converges (A detailed pseudocode of this algorithm is provided in Supplementary Figure 1). The predictions that were output from the recalibration procedure were used as the final model for predicting the risk of death from COVID-19 infection (i.e. the COVID-19 predictions).

See Supplementary Description 1 for a complete technical description of the adjustment procedure.

### Testing the adjusted outcomes

Once enough confirmed COVID-19 patients accumulated in Israel, the COVID-19 predictions were tested on the entire CHS COVID-19 patient population.

The study population was extracted on April 22^nd^, 2020, and included all patients diagnosed at least 14 days prior to the extraction date (to allow a reasonable time for the outcome to occur on the one hand, and to avoid overly reducing the sample size on the other hand). To validate this decision, empirical cumulative distribution functions of time-to-death from COVID-19 were derived using all patients in CHS’ database. These included both the crude distribution and the distribution accounting for censoring and the “competing risk” of cure, calculated using the Aalen-Johansen estimator^20^.

Patients were diagnosed using RT-PCR tests from nasal and pharyngeal swabs performed between the beginning of the outbreak in Israel and the extraction date. The outcome used was death, as identified from national registries which are updated on a daily basis.

The COVID-19 predictions were scored for discrimination using the AUROC. To assess the success of the calibration adjustment procedure, calibration was compared to that of the baseline model using calibration curves (with both the baseline and the final COVID-19 models evaluated against the COVID-19 outcome). The effects of the recalibration were also assessed via decision-curves, which present net-benefit against different decision thresholds and are used to evaluate the utility of decisions made based on the model^21^.

In addition, plots of the positive predictive value (PPV) against the sensitivity (precision-recall curve), and of the sensitivity against the percent of patients identified as high risk were drawn across different thresholds. This was done to present the sensitivity and PPV that result from all possible shares of the population that could be defined to be at high risk. These plots were also used to demonstrate the comparative performance of the binary classification that results from the American CDC criteria for defining high risk patients^4^.

### Ethical approval

Due to the urgent need for information regarding COVID-19, all data analysis related to the management of the COVID-19 pandemic in Israel (not including genetic data research and clinical trials) was exempt from a need for IRB approval (decision reference number 178368620).

### Analysis

The baseline model development was done using Python version 3.6, Anaconda version

5.1.0 and LightGBM version 2.2.3. The model performance analysis and plot creation was done in R version 3.5.2.

## Results

### Baseline Model

The characteristics of the population used for the training and testing of the final baseline model are described in Supplementary Table 2. Among this population of 1,050,000 Clalit Health Services’ (CHS) members over the age of 10 years, 11,718 (1.1%) positive outcomes were recorded over a follow-up period of one year.

The contribution and effect of the features of the baseline model for prediction of its chosen outcome, as measured by SHAP scores, are presented in Figure 1. Panel A of the figure presents the overall importance and effect of all variables in the model. Panel B presents the odds ratios across different values of three selected variables^15^.

**Figure 1.**
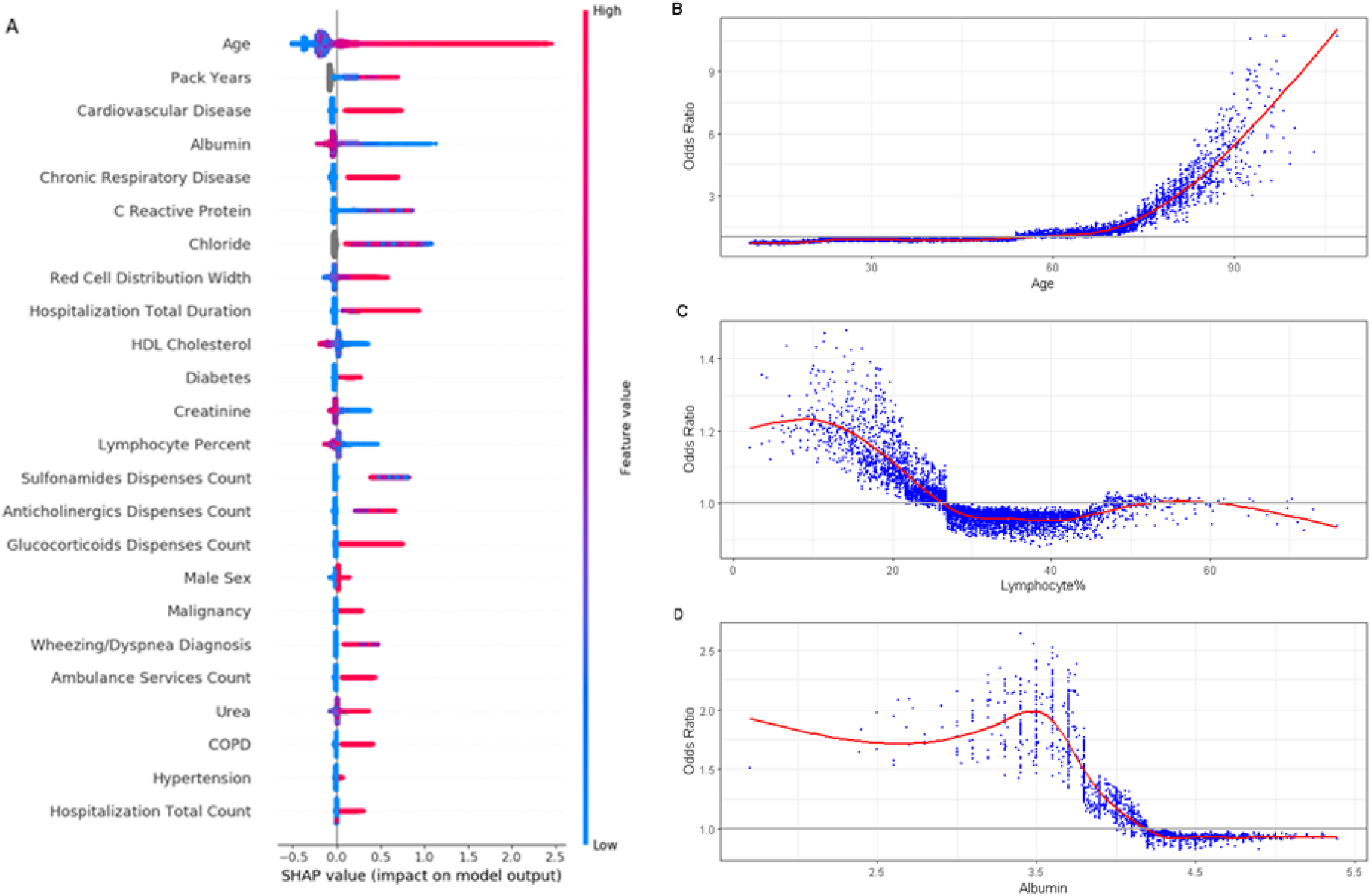
Summary and feature-specific SHAP values for the baseline model. A. A summary plot of the SHAP values for each feature. Going from top to bottom, features decrease in their overall importance (sum of SHAP values). In each feature, every point is a specific case, colors towards red represent higher values of the variable and the X-axis represents the effect the variable had on the prediction in this specific patient (increased risk to the right of the vertical grey line and decreased risk to the left of the line). B. A plot of the odds ratio for different values of age. A smoothed red line is fit to the curve and a horizontal grey line is drawn at odds ratio = 1. C. A plot of the odds ratio for different values of percent of lymphocytes in the blood. A smoothed red line is fit to the curve and a horizontal grey line is drawn at odds ratio = 1. D. A plot of the odds ratio for different values of albumin. A smoothed red line is fit to the curve and a horizontal grey line is drawn at odds ratio = 1. Abbreviations: SHAP – SHapley Additive exPlanations

Performance of the baseline prediction model on the test set is detailed in Figure 2. The AUROC was 0.820 (95% CI: 0.811-0.828), which indicates good discrimination. The calibration plot, which runs very close to the diagonal, shows excellent calibration.

**Figure 2.**
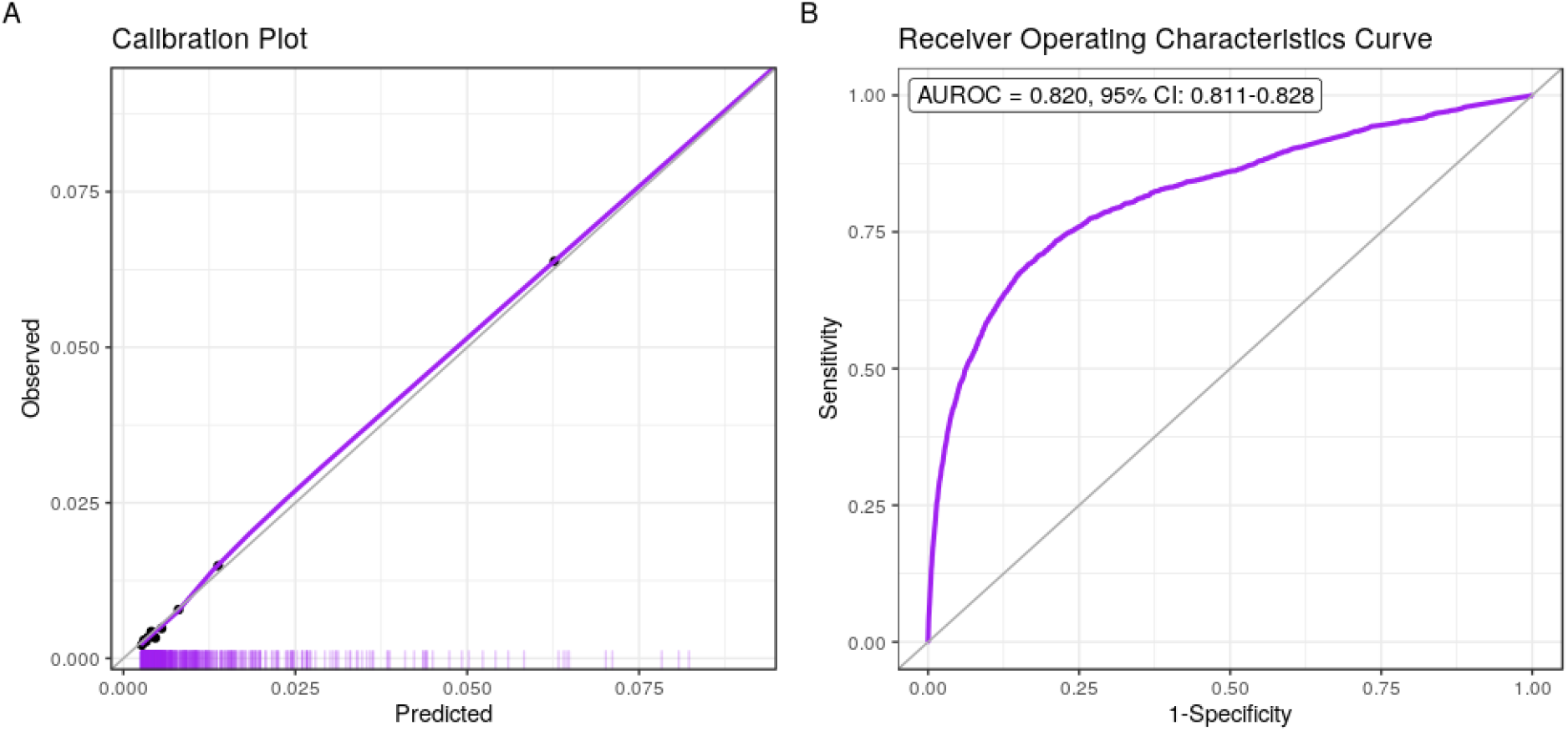
Performance charts for the baseline model. A. Calibration plot, plotting the observed outcome against the predicted probabilities. The diagonal grey line represents perfect calibration. A smoothed line is fit to the curve, and points are drawn to represent the averages in ten discretized bins. The “rug” under the plot illustrates the distribution of predictions. B. Receiver operating characteristics curve, plotting the sensitivity against one minus specificity for different values of the thresholds. The diagonal grey line represents a model with no discrimination. The area under the curve, with its 95% confidence interval, is shown on the top-left. Abbreviations: AUROC, Area Under the Receiver Operating Characteristics Curve;

### COVID-19 Complications Model

The recalibration procedure of the baseline model to the COVID-19 case fatality rates among different age and sex groups terminated after making 23 corrections. The aggregated corrections for each group are detailed in Supplementary Table 3. It is evident that the larger corrections made were for the older age groups, where the risk had to be increased substantially.

The model was deployed for large-scale use in the CHS around mid-March. When a sufficiently large local population of confirmed COVID-19 patients accumulated over the following weeks, performance of the COVID-19 predictions was tested. The last date that was allowed for confirmed cases to be included in the analysis was 14 days prior to the extraction date, thus allowing a minimum follow up period of 14 days. The empirical cumulative distribution functions for the time-of-death of all patients, both crude and adjusted for censoring and the “competing risk” of cure, is shown in Supplementary Figure 2. These figures indicate that between 79% and 88% of COVID-19 deaths occur until the 14^th^ day.

The testing population included a cohort of 3,176 COVID-19 patients, with 87 (3.1%) deaths recorded until April 22^nd^, 2020. A population table for this population is detailed in Table 1. The table shows the much higher fatality rate among the chronically ill and those of advanced age.

**Table 1.** Population characteristics table.

| **Variable** | **Outcome = No** | **Outcome = Yes** |
| --- | --- | --- |
| **Overall** | 3,089 | 87 |
| **Age group (years)** |  |  |
| 10-19 | 276 (8.9) | 0 (0.0) |
| 20-29 | 737 (23.9) | 1 (1.1) |
| 30-39 | 518 (16.8) | 1 (1.1) |
| 40-49 | 420 (13.6) | 1 (1.1) |
| 50-59 | 390 (12.6) | 1 (1.1) |
| 60-69 | 409 (13.2) | 11 (12.6) |
| 70-79 | 202 (6.5) | 20 (23.0) |
| 80-89 | 102 (3.3) | 30 (34.5) |
| 90-99 | 35 (1.1) | 22 (25.3) |
| **Sex** |  |  |
| M | 1389 (45.0) | 47 (54.0) |
| F | 1700 (55.0) | 40 (46.0) |
| **Diabetes** |  |  |
| No | 2746 (88.9) | 41 (47.1) |
| Yes | 343 (11.1) | 46 (52.9) |
| **Hypertension** |  |  |
| No | 2602 (84.2) | 28 (32.2) |
| Yes | 487 (15.8) | 59 (67.8) |
| **Cardiovascular Disease** |  |  |
| No | 2810 (91.0) | 33 (37.9) |
| Yes | 279 (9.0) | 54 (62.1) |
| **Malignancy** |  |  |
| No | 2900 (93.9) | 68 (78.2) |
| Yes | 189 (6.1) | 19 (21.8) |
| **Chronic Respiratory Disease** |  |  |
| No | 2847 (92.2) | 73 (83.9) |
| Yes | 242 (7.8) | 14 (16.1) |

Abbreviations: SD, Standard Deviation;

Discriminatory performance of the COVID-19 predictions in general and at specific thresholds is detailed in Figure 3a. The overall AUROC of the COVID-19 predictions is 0.942 (95% CI 0.916-0.960). When considering an absolute risk of 10% as the threshold, a fraction of 7% (95% CI 6%-8%) of the population is identified as having a high risk, and this high-risk group contains 69% (95% CI 59%-79%) of the patients who eventually died (sensitivity). If a patient was identified as a high risk by this cutoff, his or her probability for death was 27% (95% CI 21%-33%) (PPV). At a 5% risk threshold, a fraction of 14% (95% CI 13%-16%) of the population is found to be at high risk, with a sensitivity of 90% (95% CI 83%-96%) and PPV of 17% (95% CI 14%-21%). The CDC high-risk criteria identify 41% (95% CI 40%-43%) of the population as high risk with a sensitivity of 97% (95% CI 92%-100%) and a PPV of 6% (95% CI 5%-8%).

**Figure 3.**
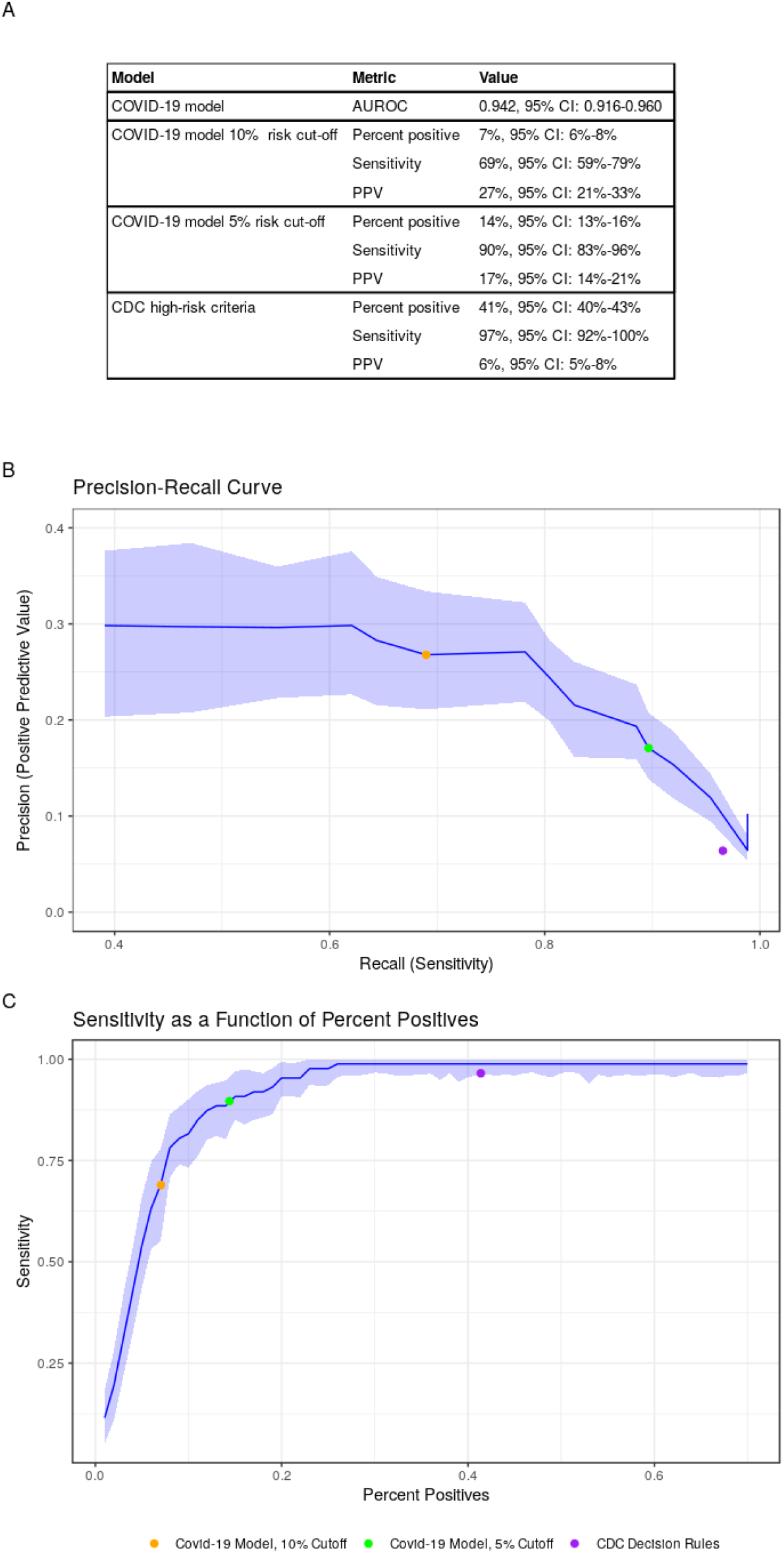
Performance charts for the COVID-19 model. A. A table specifying the performance the overall model predictions and of three classifiers. B. A plot of the positive predictive value against the sensitivity of the predictor for different thresholds. The light band around the line represents point-wise 95% confidence intervals. Only thresholds up to 15% absolute risk were plotted because of very low outcome rates in higher thresholds that which resulted in instability. The colored dots show the performance of three binary classifiers. C. A plot of the sensitivity against the percent of patients identified as high-risk for different thresholds. The light band around the line represents point-wise 95% confidence intervals. The colored dots show the performance of three binary classifiers. Abbreviations: AUROC, Area Under the Receiver Operating Characteristics Curve; CI, Confidence Interval; CDC, Centers for Disease Control and prevention; COVID-19, Corona Virus Disease 2019;

Figures 3b and 3c present plots of the COVID-19 predictions’ PPV against their sensitivity and of their sensitivity against the percent of patients identified as high risk, respectively. These plots also contain a graphical indication of the CDC criteria’s corresponding values.

A calibration plot, comparing the ability of the COVID-19 predictions and the baseline predictions in accurately evaluating the absolute risk for COVID-19 mortality, is included in Figure 4a. The figure shows that the original predictions were too low for the new outcome and that the recalibration procedure provided a marked improvement. The importance of the correction is further illustrated by decision curves (Figure 4b(. These curves, which compare the utility of the decisions that would have been made using the two models, show that decisions made using the COVID-19 predictions are superior to those made by the baseline model for the thresholds relevant for defining high-risk groups.

**Figure 4.**
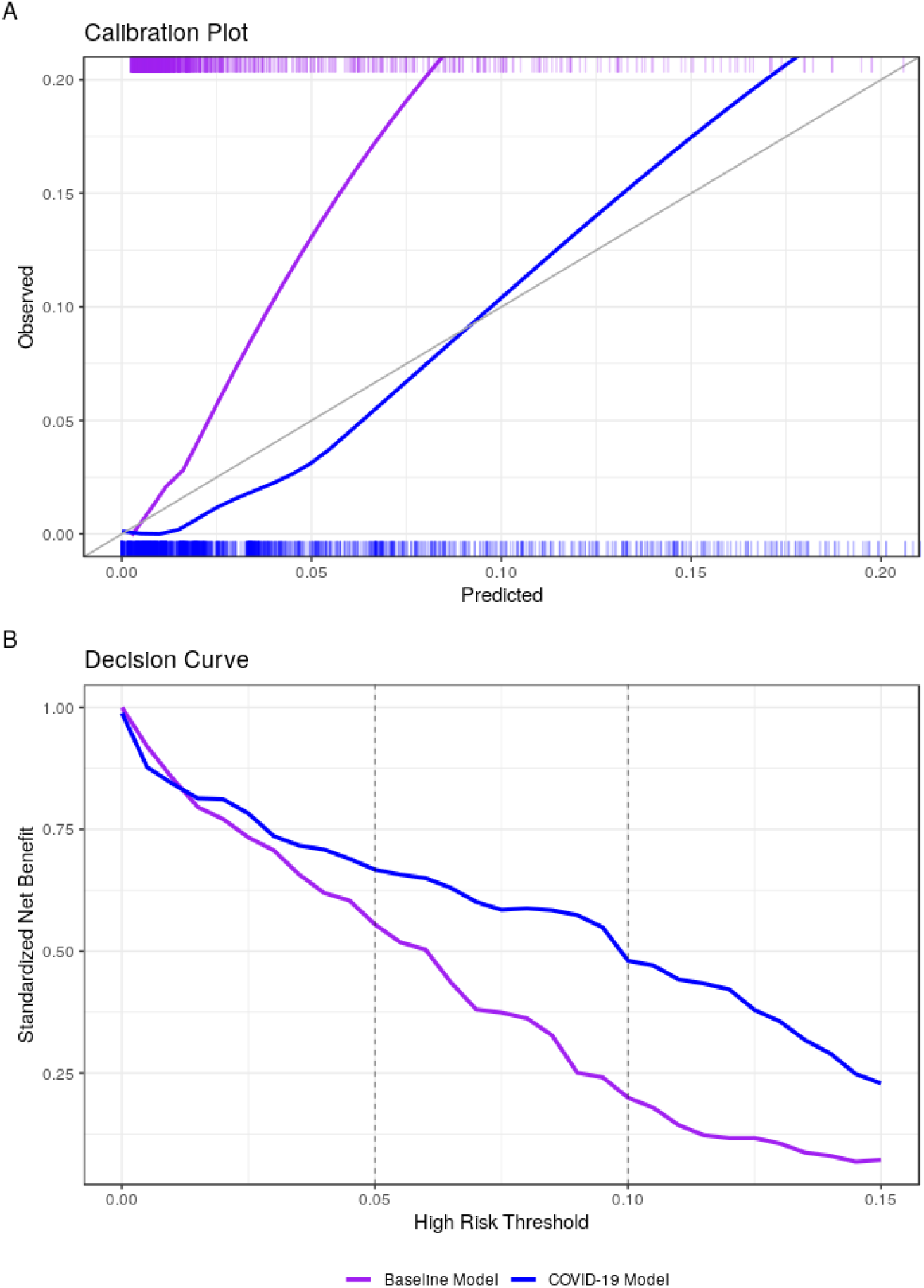
Calibration plot and decision curves comparing the COVID-19 model with the baseline model. A. Calibration plots plotting the observed outcome against the predicted probabilities of both models. The diagonal grey line represents perfect calibration. A smoothed line is fit to each curve. The “rug” above and under the plots illustrates the distribution of predictions for each model. B. The decision curve plots the standardized net benefit against different decision thresholds for both models. Net benefit is a measure of utility that calculates a weighted sum of true positives and false positives, weighted according to the threshold. Vertical dashed lines were added at relevant decision thresholds that were used in practice. Abbreviations: COVID-19, Corona Virus Disease 2019

Access details to the code required to generate the baseline model predictions, as well as the data required for the adjustment and calibration of the baseline predictions to COVID-19 CFRs, are supplied under the code availability statement.

## Discussion

### Main findings

This work presents a model for prediction of COVID-19 mortality that was developed using a hybrid approach, by calibrating the predictions from a baseline model - designed to rank the population according to the risk for severe respiratory infections or sepsis - to reported COVID-19 mortality rates. The resulting model, developed rapidly under conditions of uncertainty, entered wide use in CHS around mid-March, just before the first case of COVID-19 fatality in Israel. Since then, with the increase in the number of COVID-19 patients in Israel, some with severe outcomes, it became possible to validate its performance.

We showed that on a validation set of 3,176 COVID-19 patients (3.1% death rate), the model was highly discriminative, with an AUROC of 0.942. We also showed that the post-processing multi-calibration phase resulted in a large improvement to the calibration of the baseline model when evaluated on COVID-19 patients (Figure 4a). The improved calibration also translated to improved net benefit across the risk thresholds relevant for defining high risk groups (Figure 4b).

### The need for a risk stratification model and examples of its uses

Since its development, the model has been used for many purposes within CHS. Its first use was for prevention purposes, with an intention to notify high-risk members of their increased risk, to explain the importance of following social distancing instructions, and to provide information regarding telemedicine and other remotely accessible medical services. For that purpose, two cutoffs of high risk groups were defined: the very-high risk group (absolute predicted risk ≥ 10%, about 4.3% of the CHS population over the age of 10 years) and the high risk group (absolute predicted risk of 5-10%; 6.7% of the population). The first group received phone calls from their care providers to convey these messages personally, and both groups received a text message with similar information.

Risk stratification was also used for prioritization of COVID-19 RT-PCR tests, along with other criteria such as symptoms and potential exposure. Once tested for COVID-19, the model predictions were also used to decide on the place of treatment of confirmed cases. This was done using the following methodology: first the treating physician makes a decision regarding the preferred place for treating a new patient, based on his or her clinical condition. If it is decided that the patient can be treated outside of a hospital, the physician checks the patient’s risk group. Patients of the very-high risk group are reconsidered for hospital admission. Patients at the high-risk group can still be treated outside of a hospital, but their daily follow-up is more frequent than that of other patients.

The CDC and other authorities provided definitions of who should be considered to be at high risk^4^. There are two main issues with such binary classification. First, it does not allow the flexibility of choosing intervention cutoffs according to available resources or for interventions that are based on several risk levels. This is a problem especially when health resources are at a premium, for example when making personal phone calls by the medical staff. Second, defining a risk group by a list of risk factors (in the form of A or B or C etc.) usually results in many patients placed at the high-risk group, which is “wasteful” in its attempt to identify those who are truly at risk. This was noted in our population, where the CDC criteria define 41% of the population to be at high risk. While this group does include 97% of the death cases (sensitivity), a cutoff of 5% absolute risk by our more granular model allows defining only 14% of the population as high-risk, while still attaining a sensitivity of 90%.

### Considerations regarding model development

When first faced with the need to create a risk stratification model for COVID-19, there was no available individual level data in Israel or in the public domain. The only information available that seemed sufficiently representative (i.e. not limited to inpatient populations) was that reported by the Chinese CDC^17^, which reported CFRs for different subpopulations. Because this data cannot be translated directly to individualized risk assessment, we chose to first train a predictor on a related outcome that is available in our retrospective data and then to adjust its predicted distributions to match those COVID-19 CFRs.

Interestingly, the multi-calibration algorithm we used was borrowed from the domain of “algorithmic fairness”^22^; its intended use is to ensure calibrated and fair predictions for minority subpopulations that are underrepresented in a training set, and it is designed to run on hundreds of overlapping subpopulations (defined by many “protected” variables). In this case, however, the algorithm was used to make sure that the baseline model is calibrated for the reported CFRs of different age and sex groups. It should be noted that on top of the sex and age groups, we initially intended to calibrate the predictions to reported CFRs of populations with specific comorbidities, as any calibration target can be fed to the multi-calibration algorithm in order to further refine the predictions’ accuracy. This was eventually not done because of issues that were raised about the quality of the published comorbidity data in the Chinese report. As we could not find other reports of CFRs for different comorbidity groups in representative COVID-19 populations (as of the time of model deployment), we opted to only adjust for age and sex.

We consider this approach to provide an efficient combination between individual level data and epidemiologic reports. The resulting model can easily be further recalibrated whenever new data regarding different subpopulation CFRs becomes available, whether conditioned on a single characteristic (e.g. CFR in obese patients) or more (e.g. CFR in 50 to 70 year-old smokers).

### Interesting features

Our baseline model identified relevant features from thousands of potential candidates. Among those, the model identified seven laboratory tests as being predictive of severe respiratory illness. Interestingly, most of these tests were later found to be indicative of severe COVID-19 infection. For example, recent reports highlight lymphopenia as a marker of COVID-19 disease severity^10,23-25^. C-reactive protein (CRP) was also identified as an important feature for predicting a severe course of disease in hospitalized COVID-19 patients^10,24,26^. In addition, albumin, urea and red cell distribution width (RDW) were also mentioned as indicative of severe COVID-19 infection^26^. Lactate dehydrogenase (LDH), another variable that stood out as important is several studies^10,24-26^, was not picked up by our baseline model. These findings of overlapping risk factors further strengthen the hypothesis that the baseline model correctly identifies a physiologic tendency and relevant susceptibility for a severe COVID-19 infection.

### Comparison to previous literature

Three other models described in the literature employed a similar “baseline model” approach, with hospital admissions for respiratory disease used as a proxy for COVID-19 pneumonia. These models reported AUROC values of 0.73 to 0.81 (compared to 0.820 in our study)^10^. The models were all developed by DeCaprio et. al^27^ on a cohort of approximately 1.8 million Medicare members. Contrary to the model we report, these models were not adjusted to the COVID-19 outcome and were not evaluated on COVID-19 patients.

In addition to these, there were ten other prognostic models for predicting death, severe disease or length of admission^10^, which were trained on COVID-19 patients, almost exclusively from China. Wynants et. al^10^ determined these studies to be at high risk of bias, due to a “non-representative selection of control patients, exclusion of patients who had not experienced the event of interest by the end of the study, and high risk of model overfitting”. In addition, they criticized most of the reports for low reporting quality. It should be emphasized that due to the urgent circumstances, many of these models were published as preprints, and have not yet been subjected to a peer review process. These studies were performed in an inpatient setting, with sample sizes ranging from dozens to several hundreds and a relatively high rate of severe outcomes (some with mortality rates of over 50%). Since our work evaluates the risk for severe COVID-19 disease for the entire population, and is designed to be used prior to contracting the disease, a detailed comparison between the models is of less relevance.

### Strengths and limitations

Our work, which takes the hybrid approach of developing a population-based model that is then adjusted to COVID-19 mortality rates, has several strengths. It was conducted and reported according to the standard guidelines for prediction model reporting^11^. The baseline model was developed on a random and large sample of the CHS population, and validated on a separate test set, thus limiting the possibility for overfitting. In addition, the performance of the COVID-19 adjusted model was then evaluated on all COVID-19 confirmed CHS cases that were diagnosed at least 14 days prior to the analysis. Given that Israel has a relatively high testing rate^28^, and given that all tests and patients’ status reports are collected centrally (whether they are treated in the community or in the hospital), the possibility for selection or sampling bias is markedly reduced.

This work also has several limitations. First, the method described depends on being able to construct a discriminative model for a related outcome, for which the resulting ranking is relevant to the outcome of interest. This is usually not known accurately in advance, particularly for emerging diseases such as COVID-19. Second, the method requires subpopulation-specific CFRs that are relevant to the study population. These will usually be found in foreign populations and will need to be corrected to the local population. The correction method used in this study, using a linear probability model, is coarse, and could result in significant bias if the populations are sufficiently different. Third, patients that were diagnosed later in the follow-up period had a shorter follow-up time (with a minimum of 14 days) from the time-of-diagnosis to the time the outcome of death was ascertained. As our analysis shows, a 14 days of follow-up has the potential to miss up to 20% more deaths.

Last and most important, without individual level data, there is no way to test the resulting predictions. As a result, a poorly performing model could find its way into clinical use at a critical time. This can be counteracted, at least partially, by evaluating the resulting model and predictions manually, using clinical domain knowledge, as was performed in the CHS prior to model deployment. This also emphasizes the importance of validating the model on individual level data as soon as such data becomes available.

## Conclusions

In this work we described the development and use of a prediction model for a novel infection, when individual level data is not yet available. We found that even in the midst of a pandemic, shrouded in epidemiologic “fog of war”, it was possible to provide a useful prediction model for COVID-19 with good discrimination and calibration. It appears that the choice of a related outcome was able to provide a proper ranking of the population, and the multi-calibration approach was useful for integration of published epidemiological data into the model. We also demonstrated that this approach is more efficient in identifying high-risk individuals compared to the current CDC definitions.

The methodology described here can be used in other populations lacking individual COVID-19 data and in future similar circumstances for other emerging diseases or situations in which an outcome of interest is not yet available in the local data. As a healthcare organization, we consider risk stratification tools as important for both proactive prevention measures and for care decisions regarding confirmed patients. This need becomes even more critical when health systems are facing extreme loads and there is a need to properly assign available resources.

## Data Availability

Data availability
Access to the data used for this study can be made available upon request, subject to an internal review by N.B. and N.D. to ensure that participant privacy is protected, and subject to completion of a data sharing agreement, approval from the institutional review board of Clalit Health Services and institutional guidelines and in accordance with the current data sharing guidelines of Clalit Health Services and Israeli law. Pending the aforementioned approvals, data sharing will be made in a secure setting, on a per-case-specific manner, as defined by the chief information security officer of Clalit Health Services. Please submit such requests to N.D.
Code availability
The analytic code required to produce the COVID-19 model predictions is available at: https://github.com/clalitresearch/COVID-19-Model

https://github.com/clalitresearch/COVID-19-Model

## Acknowledgements

We want to thank Adi Berliner, Shay Ben-Shahar, Anna Kuperberg, Reut Ohana, Nurit Man, Irena Livshitz, Alon Schwartz, Nir Shahar and Ilana Roitman for contributing to the model’s implementation. We want to thank Mark Katz, Anna Kuperberg, Morton Leibowitz and Oren Oster for helping with the outcome definition. We want to thank Shay Perchik, Michael Lischinsky and Galit Shaham for helping with Quality Assurance. We want to thank Ilan Gofer for organizing the data.

## Contributions

N.B., D.R., R.B. and N.D. conceived and designed the study. N.B., D.R., A.A, J.L., U.F. and N.D. participated in data extraction and analysis. G.Y., D.G., S.S., J.S., G.N.R and U.S. participated in the conception and creation of the model adjustment process. N.B. and N.D. wrote the manuscript. All authors critically revised the manuscript. R.B and N.D. supervised the entire study process.

## Competing interests

All authors have no competing interests to disclose.

## Funding Statement

G.N.R. reports grants from Israel Science Foundation, grants from Israel-US Binational Science Foundation, grants from European Research Council, grants from Amazon Research Award during the conduct of the study. U.S. reports personal fees from K-health, outside the submitted work.

## Data availability

Access to the data used for this study can be made available upon request, subject to an internal review by N.B. and N.D. to ensure that participant privacy is protected, and subject to completion of a data sharing agreement, approval from the institutional review board of Clalit Health Services and institutional guidelines and in accordance with the current data sharing guidelines of Clalit Health Services and Israeli law. Pending the aforementioned approvals, data sharing will be made in a secure setting, on a per-case-specific manner, as defined by the chief information security officer of Clalit Health Services. Please submit such requests to N.D.

## Code availability

The analytic code required to produce the COVID-19 model predictions is available at: https://github.com/clalitresearch/COVID-19-Model

## Supplementary Material

**Supplementary Figure 1.**
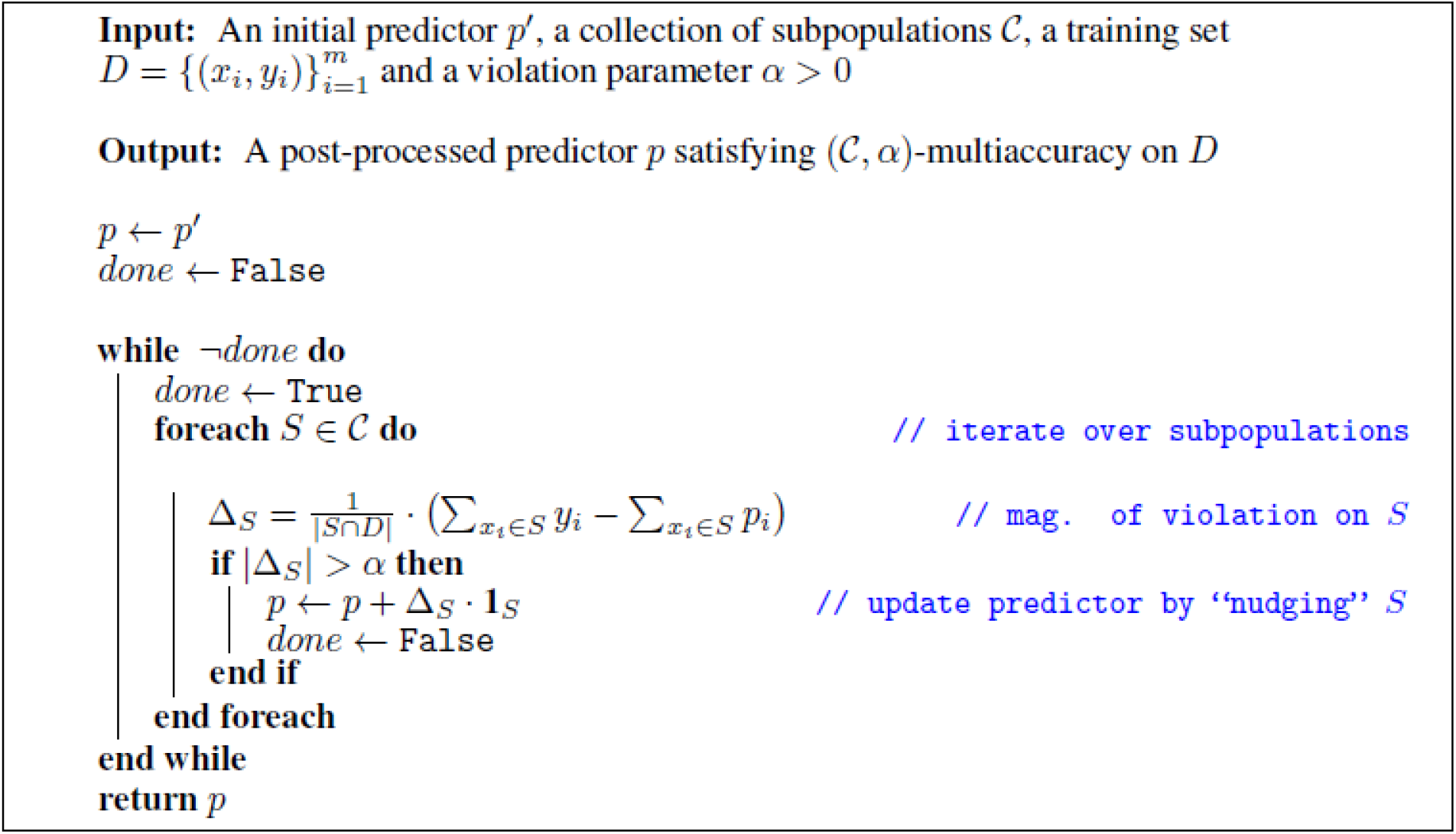
Pseudo-code of the “multi-calibration” algorithm. Detailed pseudo-code of the multi-calibration algorithm, as described in Hebert-Johnson et al.^12^

**Supplementary Figure 2.**
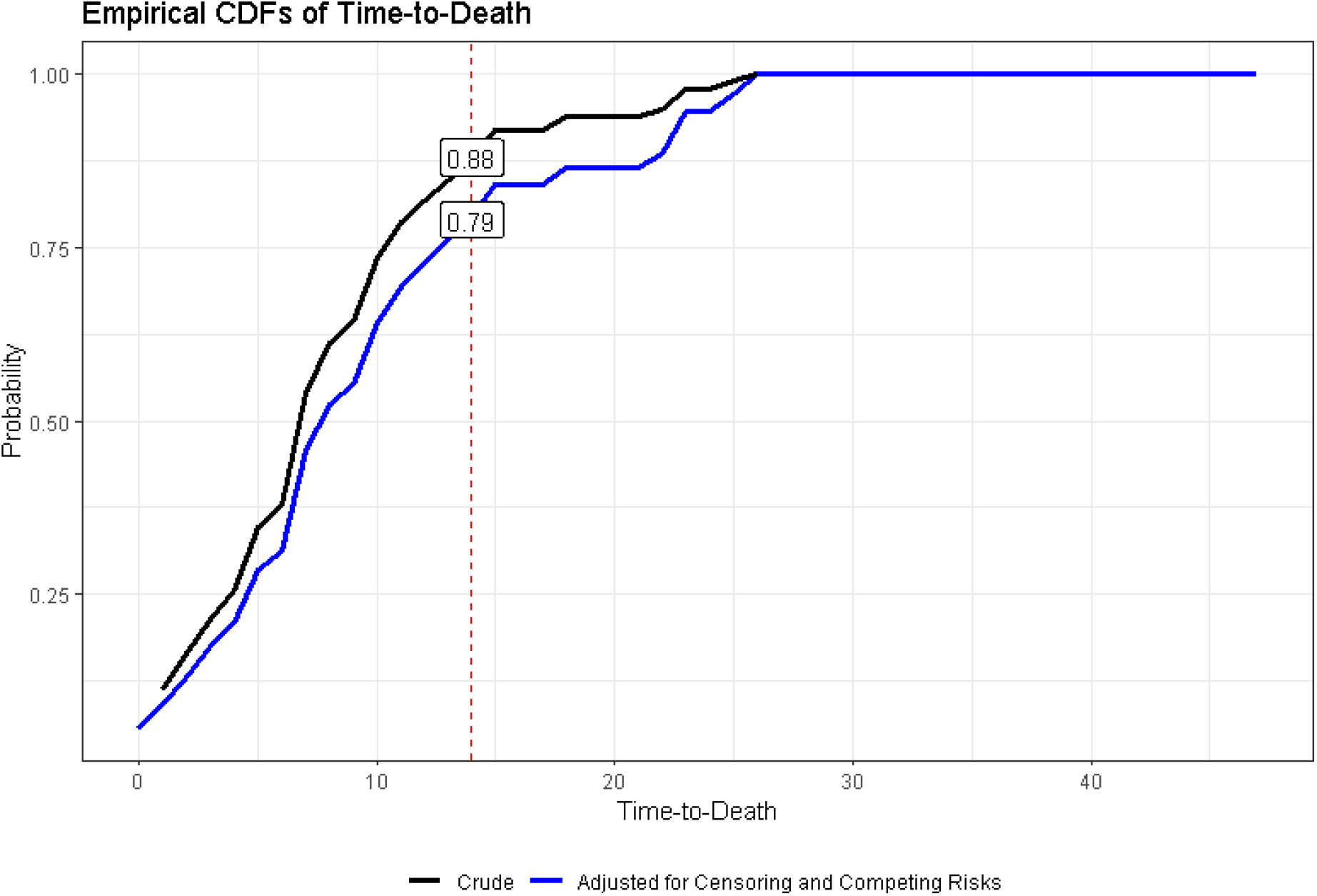
Cumulative Distribution Functions of time-to-death in CHS’ database. Empirical cumulative distribution functions for time-to-death of all COVID-19 patients in CHS’ database. The black line shows the crude distribution. The blue line shows the cumulative incidence as calculated after accounting for censoring the “competing risk” of cure, derived using the Aalen-Johansen estimator^20^.

### Supplementary Description 1 – Technical description of adjustment procedure

Purpose: to adjust predictions to external one-way conditional probabilities. Inputs:

- Baseline predictions for another outcome on the entire population of interest, P(Y_old_|X), conditioned on as many independent variables as wanted.
- Published one-way conditional probabilities for the outcome of interest, from an external population, conditional on n variables. P_external_(Y_new_|X_i), I = 1..n
- A co-occurrence matrix M of size n_x_n, where M_i,j_ = P_external_(X_j_|X_i_) in the population on which the one-way conditional probabilities were published
- Rates of the different population characteristics in the local population, P_local_(X_i), I = 1..n

Steps:

1. <u>Solving for the coefficients of the linear probability model</u> This is done by deriving the coefficients for each independent variable in a linear probability model. That is, the model is P(Y) = w_1_P(X_1_) + … + w_n_P(X_n_), and the coefficients are solved for by solving a system of n equations:

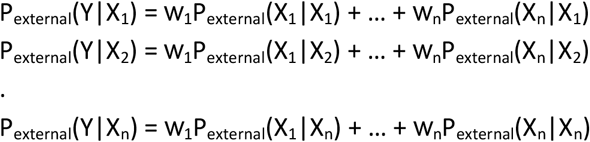 Where P(X_i_|X_j_) is the i,j entry of the M matrix.
2. <u>Adjust external one-way conditionals to the local population</u> With the coefficients in hand, rate for the local population are calculated using the local probabilities of the independent variables,

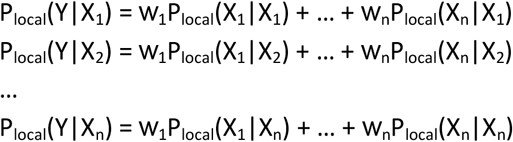
3. <u>Recalibrate the predictions for the baseline outcome to the outcome rates of the new outcome</u>

With the calculated local outcome rates in hand, the baseline predictions are adjusted using the multi-accuracy algorithm depicted in Supplemental Figure 1.

When the process terminates, we have P(Y_new_|X) for the entire population of interest.

**Supplementary Table 1.** Variable and outcome definitions.

| Variable Definitions | Category | Units | Time Frame (prior to index date) | Details | Mechanism of feature selection |
| --- | --- | --- | --- | --- | --- |
| Outcome | Diagnoses | No / Yes |  | ICD9 codes 48[0-8]*, 46[0-6]*, 490*, 510.9*, 511.0, 511.89, 518.82, 785.52 or positive influenza PCR | NA |
| Age | Demographics | Years | Current | Age in full years | From the top-features list |
| Sex | Demographics | Male / Female | Current |  | Added manually |
| Pack years | Clinical Covariates | Count | 1 year | #years smoked X average number of packs per year | From the top-features list |
| COPD | Diagnoses | No / Yes | Ever | As per Clalit's chronic disease registry | From the top-features list |
| Number of wheezing / dyspnea diagnoses | Diagnoses | Count | 1 year | ICD9: 786.0 | From the top-features list |
| Albumin | Labs | g/dl | 1 year |  | From the top-features list |
| Red cell distribution width | Labs | % | 1 year |  | From the top-features list |
| C-Reactive Peptide | Labs | mcg/ml | 1 year |  | From the top-features list |
| Urea | Labs | mg/dl | 1 year |  | From the top-features list |
| Lymphocyte | Labs | % | 1 year |  | From the top-features list |
| Chloride | Labs | mEq/L | 1 year |  | From the top-features list |
| Creatinine | Labs | mg/dl | 1 year |  | From the top-features list |
| High Density Lipoprotein | Labs | mg/dl | 1 year |  | From the top-features list |
| Duration of hospitalizations | Healthcare Utilization | Count | 1 year |  | From the top-features list |
| Count of hospitalizations | Healthcare Utilization | Count | 1 year |  | From the top-features list |
| Count of ambulance rides | Healthcare Utilization | Count | 1 year |  | From the top-features list |
| Count of Sulfonamide dispenses | Drugs | Count | 1 year | ATC4: C03CA | From the top-features list |
| Count of Anti-cholinergic dispenses | Drugs | Count | 1 year | ATC4: R03BB | From the top-features list |
| Count of Glucocorticoid dispenses | Drugs | Count | 1 year | ATC4: H02AB | From the top-features list |
| Chronic Respiratory Disease | Diagnoses | No / Yes | Ever | As per Clalit's chronic disease registry | Added manually |
| Cardiovascular Disease | Diagnoses | No / Yes | Ever | As per Clalit's chronic disease registry | Added manually |
| Diabetes | Diagnoses | No / Yes | Ever | As per Clalit's chronic disease registry | Added manually |
| Malignancy | Diagnoses | No / Yes | Ever | As per Clalit's chronic disease registry | Added manually |
| Hypertension | Diagnoses | No / Yes | Ever | As per Clalit's chronic disease registry | Added manually |

Caption: A list of the variables used in the model, including their type, units, time frame of extraction, definitions and how they were selected.
Abbreviations: COPD, Chronic Obstructive Pulmonary Disease; g, gram; mcg, microgram; mEq, mili-equivalent; ml, milliliter; dl, deciliter; L, liter; PCR, Polymerase Chain Reaction; ICD9, International Classification of Disease, 9<sup>th</sup> revision; ATC4, Anatomical Therapeutic Chemical (ATC) Classification System, 4<sup>th</sup> level;

**Supplementary Table 2.**
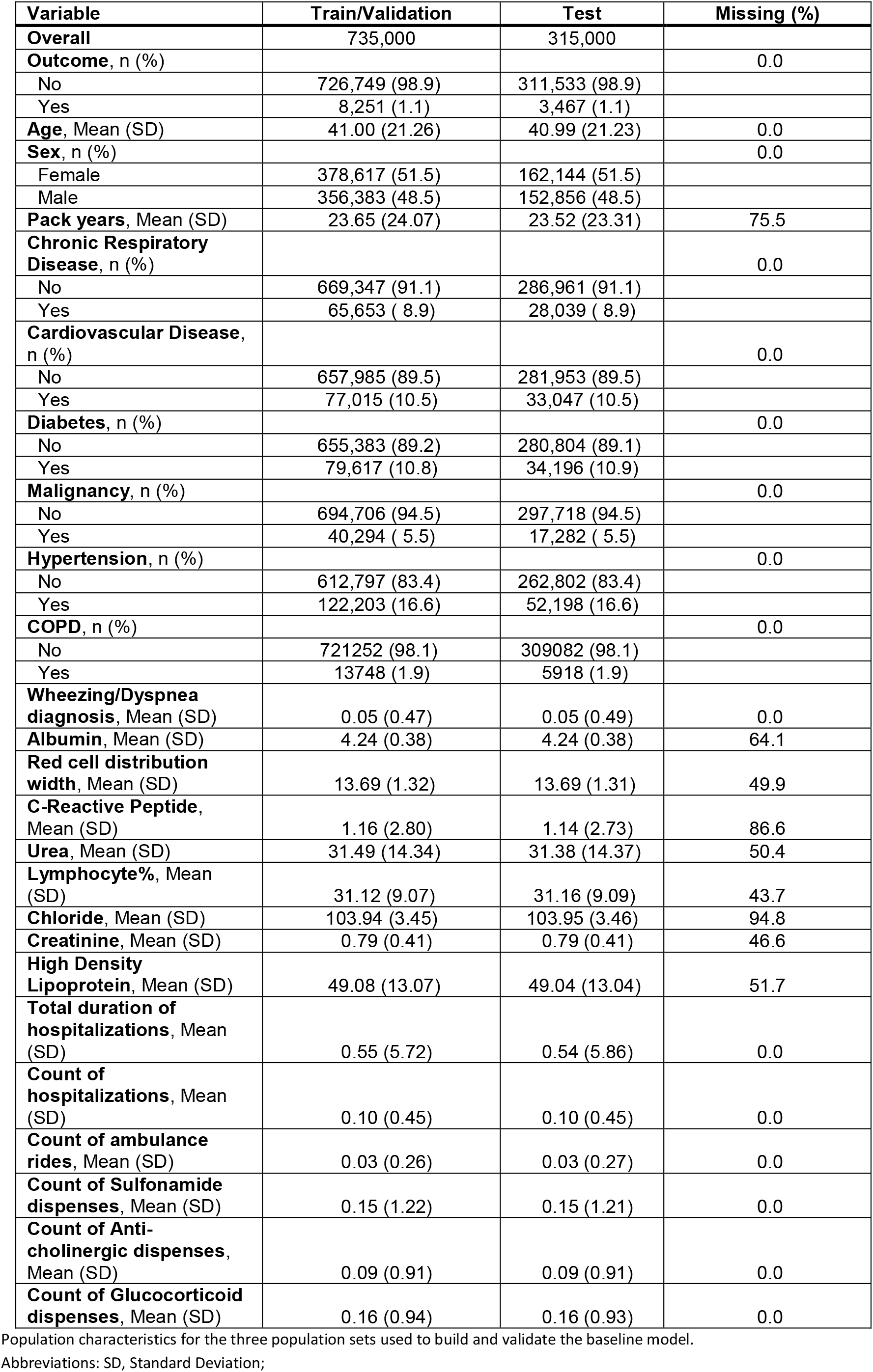
Population table for baseline model.

| Variable | Train/Validation | Test | Missing (%) |
| --- | --- | --- | --- |
| **Overall** | 735,000 | 315,000 |  |
| **Outcome, n (%)** |  |  | 0.0 |
| No | 726,749 (98.9) | 311,533 (98.9) |  |
| Yes | 8,251 (1.1) | 3,467 (1.1) |  |
| **Age, Mean (SD)** | 41.00 (21.26) | 40.99 (21.23) | 0.0 |
| **Sex, n (%)** |  |  | 0.0 |
| Female | 378,617 (51.5) | 162,144 (51.5) |  |
| Male | 356,383 (48.5) | 152,856 (48.5) |  |
| **Pack years, Mean (SD)** | 23.65 (24.07) | 23.52 (23.31) | 75.5 |
| **Chronic Respiratory Disease, n (%)** |  |  | 0.0 |
| No | 669,347 (91.1) | 286,961 (91.1) |  |
| Yes | 65,653 ( 8.9) | 28,039 ( 8.9) |  |
| **Cardiovascular Disease, n (%)** |  |  | 0.0 |
| No | 657,985 (89.5) | 281,953 (89.5) |  |
| Yes | 77,015 (10.5) | 33,047 (10.5) |  |
| **Diabetes, n (%)** |  |  | 0.0 |
| No | 655,383 (89.2) | 280,804 (89.1) |  |
| Yes | 79,617 (10.8) | 34,196 (10.9) |  |
| **Malignancy, n (%)** |  |  | 0.0 |
| No | 694,706 (94.5) | 297,718 (94.5) |  |
| Yes | 40,294 ( 5.5) | 17,282 ( 5.5) |  |
| **Hypertension, n (%)** |  |  | 0.0 |
| No | 612,797 (83.4) | 262,802 (83.4) |  |
| Yes | 122,203 (16.6) | 52,198 (16.6) |  |
| **COPD, n (%)** |  |  | 0.0 |
| No | 721252 (98.1) | 309082 (98.1) |  |
| Yes | 13748 (1.9) | 5918 (1.9) |  |
| **Wheezing/Dyspnea diagnosis, Mean (SD)** | 0.05 (0.47) | 0.05 (0.49) | 0.0 |
| **Albumin, Mean (SD)** | 4.24 (0.38) | 4.24 (0.38) | 64.1 |
| **Red cell distribution width, Mean (SD)** | 13.69 (1.32) | 13.69 (1.31) | 49.9 |
| **C-Reactive Peptide, Mean (SD)** | 1.16 (2.80) | 1.14 (2.73) | 86.6 |
| **Urea, Mean (SD)** | 31.49 (14.34) | 31.38 (14.37) | 50.4 |
| **Lymphocyte%, Mean (SD)** | 31.12 (9.07) | 31.16 (9.09) | 43.7 |
| **Chloride, Mean (SD)** | 103.94 (3.45) | 103.95 (3.46) | 94.8 |
| **Creatinine, Mean (SD)** | 0.79 (0.41) | 0.79 (0.41) | 46.6 |
| **High Density Lipoprotein, Mean (SD)** | 49.08 (13.07) | 49.04 (13.04) | 51.7 |
| **Total duration of hospitalizations, Mean (SD)** | 0.55 (5.72) | 0.54 (5.86) | 0.0 |
| **Count of hospitalizations, Mean (SD)** | 0.10 (0.45) | 0.10 (0.45) | 0.0 |
| **Count of ambulance rides, Mean (SD)** | 0.03 (0.26) | 0.03 (0.27) | 0.0 |
| **Count of Sulfonamide dispenses, Mean (SD)** | 0.15 (1.22) | 0.15 (1.21) | 0.0 |
| **Count of Anti-cholinergic dispenses, Mean (SD)** | 0.09 (0.91) | 0.09 (0.91) | 0.0 |
| **Count of Glucocorticoid dispenses, Mean (SD)** | 0.16 (0.94) | 0.16 (0.93) | 0.0 |

Population characteristics for the three population sets used to build and validate the baseline model.
Abbreviations: SD, Standard Deviation;

**Supplementary Table 3.**
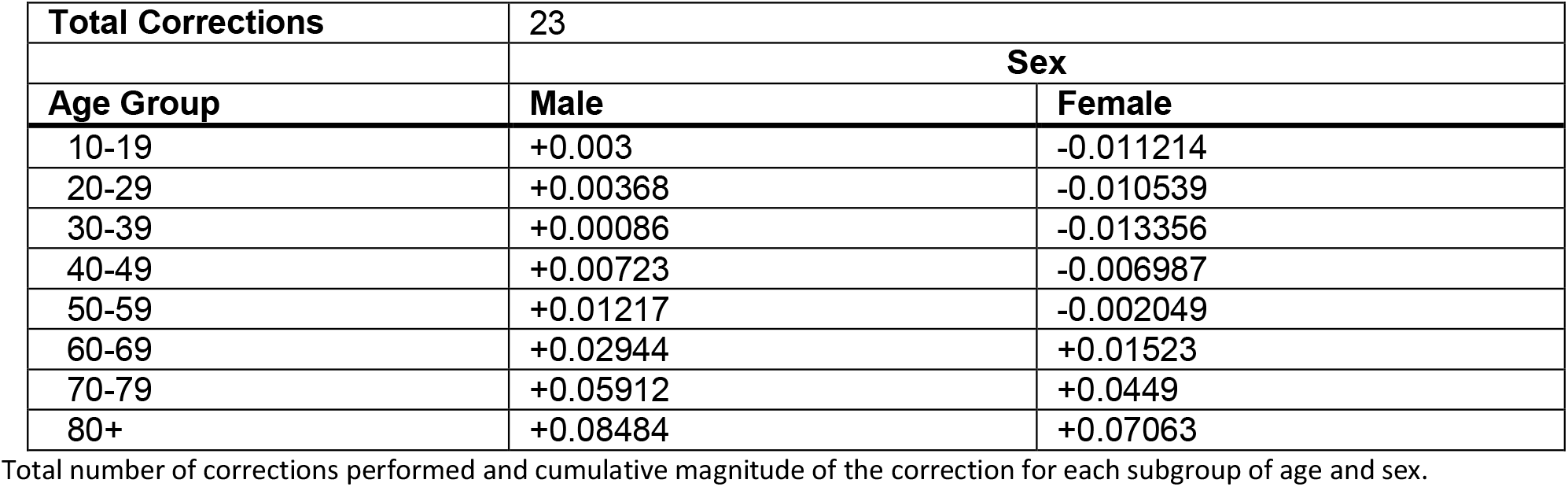
Correction table for the “multi-calibration” recalibration procedure.

|  |  |  |
| --- | --- | --- |
| **Total Corrections** | 23 |  |
|  | **Sex** |  |
| **Age Group** | **Male** | **Female** |
| 10-19 | +0.003 | -0.011214 |
| 20-29 | +0.00368 | -0.010539 |
| 30-39 | +0.00086 | -0.013356 |
| 40-49 | +0.00723 | -0.006987 |
| 50-59 | +0.01217 | -0.002049 |
| 60-69 | +0.02944 | +0.01523 |
| 70-79 | +0.05912 | +0.0449 |
| 80+ | +0.08484 | +0.07063 |

Total number of corrections performed and cumulative magnitude of the correction for each subgroup of age and sex.

## Notes

### Competing Interest Statement

The authors have declared no competing interest.

